# Real-time wearable detection of respiratory infections in pediatric transplant households

**DOI:** 10.64898/2026.10.09.26365132

**Authors:** Caleb Mayer, Eric Hurwitz, Tanya Bhatia, Annie Chung, Divya Gandla, Bryanna Domenick, Esa Räsänen, Nik Nair, Ekanath Srihari Rangan, Humera Ahmed, Chelsie Brathwaite, Mauricio Arcila-Meso, Hui Gao, Kleio-Maria Verrou, Vincent Mauduit, Maureen B. Parenti, Elizabeth Rand, Amit A. Shah, Cameron Myhrvold, Chan Wang, Sandra Amaral, Matthew J. O’Connor, Michael P. Snyder, Brendan Keating

## Abstract

Consumer-grade wearables can continuously and longitudinally measure health signals such as heart rate (HR), activity, and sleep. While changes in these signals have been implicated in the early detection of infection (e.g., influenza, SARS-CoV-2) and real-time alerting systems have been developed, evidence from prospective deployment in pediatric transplant households remains limited. Here, we study the application of a resting heart rate (RHR)-based real-time alerting algorithm to detect respiratory virus infection in a cohort of solid organ transplant recipients and their household members (n=146) in a Phase IV clinical trial (NCT06161454). Reliable and efficient alerting for infection events may be particularly relevant for this population, enabling isolation, clinical management, and/or treatment. Of 31 antigen-test outcomes, nearly all (29/31, 93.5%) correspond to either influenza, COVID, or other patient-reported symptoms. We describe RHR, HR, activity, and sleep/circadian rhythm features around clinical alarm events, identifying candidate features for further investigation and algorithm improvement.

## INTRODUCTION

Early detection of respiratory viral infections remains an important clinical and public health goal ^1–3^. For infections such as influenza and COVID-19, timely recognition at either an individual or clinical level can enable testing, isolation, clinical monitoring, and antiviral treatment ^4^. Although influenza tends to be self-limiting in healthy adults, severe infection can result in prolonged hospitalization, intensive care unit (ICU) admission, and death, particularly among children, older adults, and immunocompromised individuals ^5^. Current healthcare strategies for infection are largely reactive, depending on patient recognition of symptoms and subsequent testing or clinical care. This creates a lag between biological infection onset and clinical action, as measurable physiological perturbations can precede symptom onset. Approaches that detect these early deviations from an individual’s baseline may be able to identify infection during a window when personal and clinical action is most useful.

Early infection detection is particularly relevant for solid organ transplant populations ^6–9^. Transplant recipients are clinically vulnerable to respiratory viral infections, due to immunosuppression, underlying disease, and post-transplant clinical complexity ^6,10,11^. Respiratory viruses such as influenza and SARS-CoV-2 can lead to more severe acute illness, hospitalization, and disruption of transplant care, and complications from infection after transplant are a major driver of mortality in these cohorts ^12–14^. In prospective multicenter studies of adult solid-organ transplant recipients with clinically managed influenza, approximately 66-71% required hospitalization, more than 30% developed pneumonia, 11-16% required ICU admission, and approximately 4% died ^15,16^. Thus, delays in diagnosis and clinical intervention may be particularly consequential. Early infection identification may be especially valuable in pediatric recipients, among whom respiratory viral infections are common and can result in substantial hospitalization, morbidity, and transplant-related complications ^17^. In addition to the pediatric transplant patient themself, infection in surrounding household members increases transmission risk. Following an influenza infection in one household member, reported transmission risks to household contacts have reached approximately 38%, with secondary infections typically occurring within several days of illness onset in the original case ^18^. Monitoring the transplant recipient alone may therefore miss important opportunities for early family-level risk detection and preventive action ^12,19^.

Wearables enable the collection of dense, longitudinal physiological and behavioral data, including heart rate (HR), resting heart rate (RHR), heart rate variability (HRV), activity, and sleep, in real-world settings. These data support tracking health over time through within-person monitoring, where high data density allows each individual to serve as their own baseline. Smartwatches and other wearables thus provide clinically relevant information that complements episodic in-clinic assessments and support remote monitoring, risk identification, and earlier recognition of health status changes. Prior work has identified alterations in wearable features before or around infection-related symptom onset ^20–26^. Retrospective analyses have shown deviations in RHR, the relationship between HR and activity, and circadian features around infection onset ^20,21^. Indeed, RHR is one of the more consistently available wearable physiological measures, and captures systemic autonomic and inflammatory responses that accompany viral infection. Real-time alerting systems based on these and other deviations in wearable features have extended this work ^22,27,28^. Notably, the NightSignal algorithm enables real-time detection of infections based on sustained deviations in nighttime RHR ^29^. NightSignal leverages repeated wearable measurements to establish personalized baselines rather than fixed population thresholds, and generates a daily alert status (red, yellow, or green).

In this study, we prospectively deployed the NightSignal algorithm in a cohort of pediatric solid organ transplant recipients (SOTR) and their household members. Participants uploaded their wearable data, while the study team tracked the resulting alerts, contacted participants following repeated red alerts, and initiated at-home antigen testing and baloxavir treatment when deemed clinically appropriate as per study protocol. We evaluate the prospective deployment of this wearable alerting system in pediatric transplant households. We describe the cohort and wearable data availability, the frequency and timing of NightSignal alerts, and antigen testing outcomes following alerts and associated with recorded clinical alarms. We further examine wearable trajectories around infection-related alerts/alarms and antigen testing, comparing influenza-positive, COVID-19-positive, and antigen-negative cases. Finally, we quantify patterns in RHR, the HR/activity relationship, and sleep and circadian rhythm disruption preceding and during alarm events.

## RESULTS

### Wearable cohort characteristics and data availability

The wearable analysis cohort consisted of 146 participants with at least one day of HR data; 43 pediatric transplant patients and 103 household members (**Figure 1; Table 1**). Participants had a median age of 31.1 years (IQR = [20.3,50.2]), with transplant patients being younger than household members (median age 20.1 years versus 44.6 years, respectively). The cohort was 61.6% female overall, and 91.1% of participants were associated with liver or kidney transplant households. Wearable data availability varied across participants but supported longitudinal assessment of HR over extended follow-up periods, with a median of 71.5 days of HR data per participant. Household members had greater data availability than patients, with a median of 78.0 days compared with 55.0 days among patients, although this difference was not statistically significant after accounting for household clustering (household-member/patient geometric-mean ratio = 1.34, 95% CI [0.82, 2.18], P = 0.241). Participant-level average daily minimum RHR was available for 106 participants and was higher among patients than household members, with a median of 65.6 bpm in patients compared with 58.6 bpm in household members (mean difference, patient minus household member = 7.54 bpm, 95% CI [3.81, 11.28], P < 0.001). See **Table 1** and **Figure 1** for further information on the cohort.

**Table 1:**
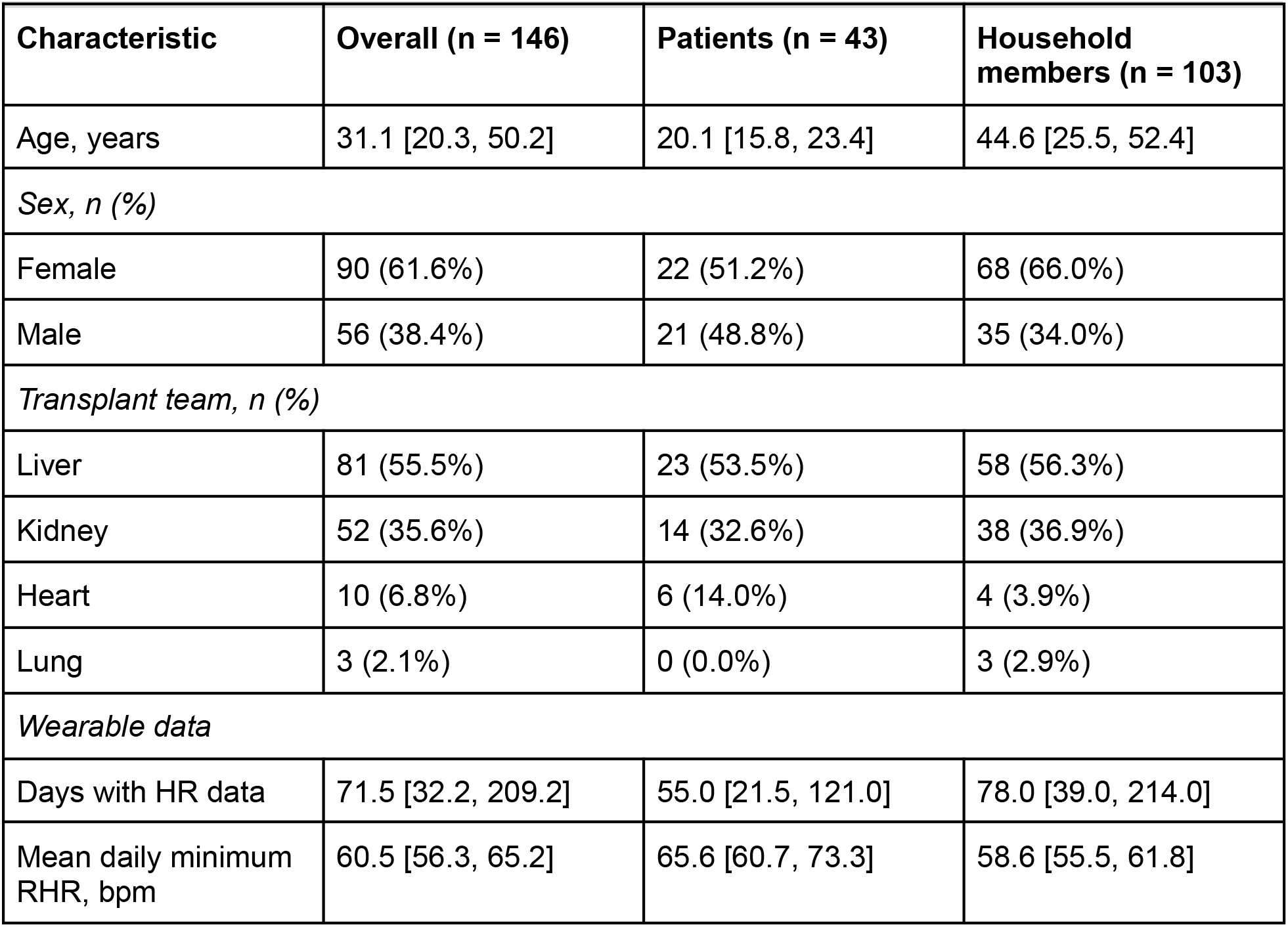
Cohort characteristics and wearable data availability. Characteristics are shown for the overall wearable cohort and stratified by participant type. Values are median [interquartile range] or n (%) unless otherwise indicated. Mean daily minimum RHR was available for 106 participants: 33 patients and 73 household members. Transplant team denotes the clinical transplant program associated with the participant’s household.

| Characteristic | Overall (n = 146) | Patients (n = 43) | Household members (n = 103) |
| --- | --- | --- | --- |
| Age, years | 31.1 [20.3, 50.2] | 20.1 [15.8, 23.4] | 44.6 [25.5, 52.4] |
| *Sex, n (%)* |  |  |  |
| Female | 90 (61.6%) | 22 (51.2%) | 68 (66.0%) |
| Male | 56 (38.4%) | 21 (48.8%) | 35 (34.0%) |
| *Transplant team, n (%)* |  |  |  |
| Liver | 81 (55.5%) | 23 (53.5%) | 58 (56.3%) |
| Kidney | 52 (35.6%) | 14 (32.6%) | 38 (36.9%) |
| Heart | 10 (6.8%) | 6 (14.0%) | 4 (3.9%) |
| Lung | 3 (2.1%) | 0 (0.0%) | 3 (2.9%) |
| *Wearable data* |  |  |  |
| Days with HR data | 71.5 [32.2, 209.2] | 55.0 [21.5, 121.0] | 78.0 [39.0, 214.0] |
| Mean daily minimum RHR, bpm | 60.5 [56.3, 65.2] | 65.6 [60.7, 73.3] | 58.6 [55.5, 61.8] |

**Figure 1.**
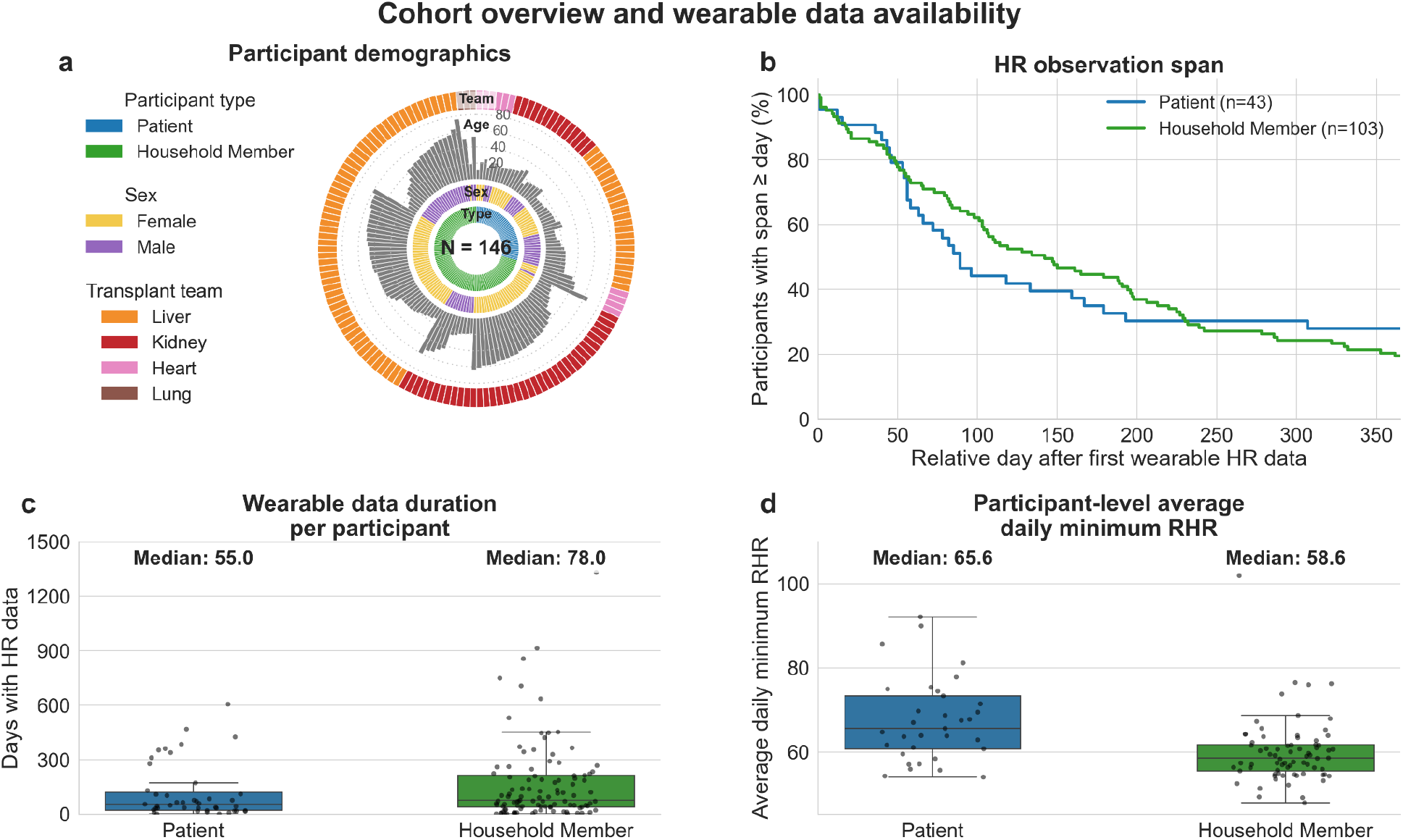
Cohort overview and wearable data availability. **a**, Distribution of participants included in the wearable analysis. The outermost level shows the transplant team (Liver, Kidney, Heart, or Lung), the next level shows the Age, the next level the Sex, and the innermost level the Participant type (Patient or Household Member). **b**, Percentage of participants whose HR data span (last minus first observed date) reached each relative day, shown through 365 days using fixed denominators of 43 patients (blue) and 103 household members (green). **c**, Number of days with available wearable HR data per participant, stratified by participant group. Boxes show the median and interquartile range, and points represent individual participants. Median values are shown above each group. **d**, Average daily minimum RHR at the participant level, calculated from available daily minimum RHR values and stratified by participant group. Boxes show the median and interquartile range, and points represent individual participants. Median values are shown above each group.

### Red alert rates were higher in the pre-alarm window

From the wearable cohort and associated clinical records, 31 antigen-test outcomes were identified (see Methods). The 31 antigen-test outcome events were contributed by 21 participants from 13 households, with several participants contributing more than one event (**Supplemental Figure 1**). Thirty records from 20 participants had a recorded wearable identifier available for linkage. Among these events with antigen testing results, 18 of 31 (58.1%) were negative for both influenza and SARS-CoV-2, 7 (22.6%) were SARS-CoV-2 positive and influenza negative, and 6 (19.4%) were influenza positive and SARS-CoV-2 negative (**Figure 2a**). Of the 18 events corresponding to a negative test for influenza and SARS-CoV-2, there were 20 recorded symptom-category reports, which consisted of cold or respiratory symptoms (9/20, 45.0%), other symptoms or illness (4/20, 20.0%), gastrointestinal symptoms (3/20, 15.0%), no symptoms (2/20, 10.0%), mental health-related symptoms (1/20, 5.0%), and menstruation-related symptoms (1/20, 5.0%; **Figure 2b**). These results indicate that the wearable alerts identified episodes including both confirmed viral infection and antigen-negative events associated with other illness or symptoms.

**Figure 2.**
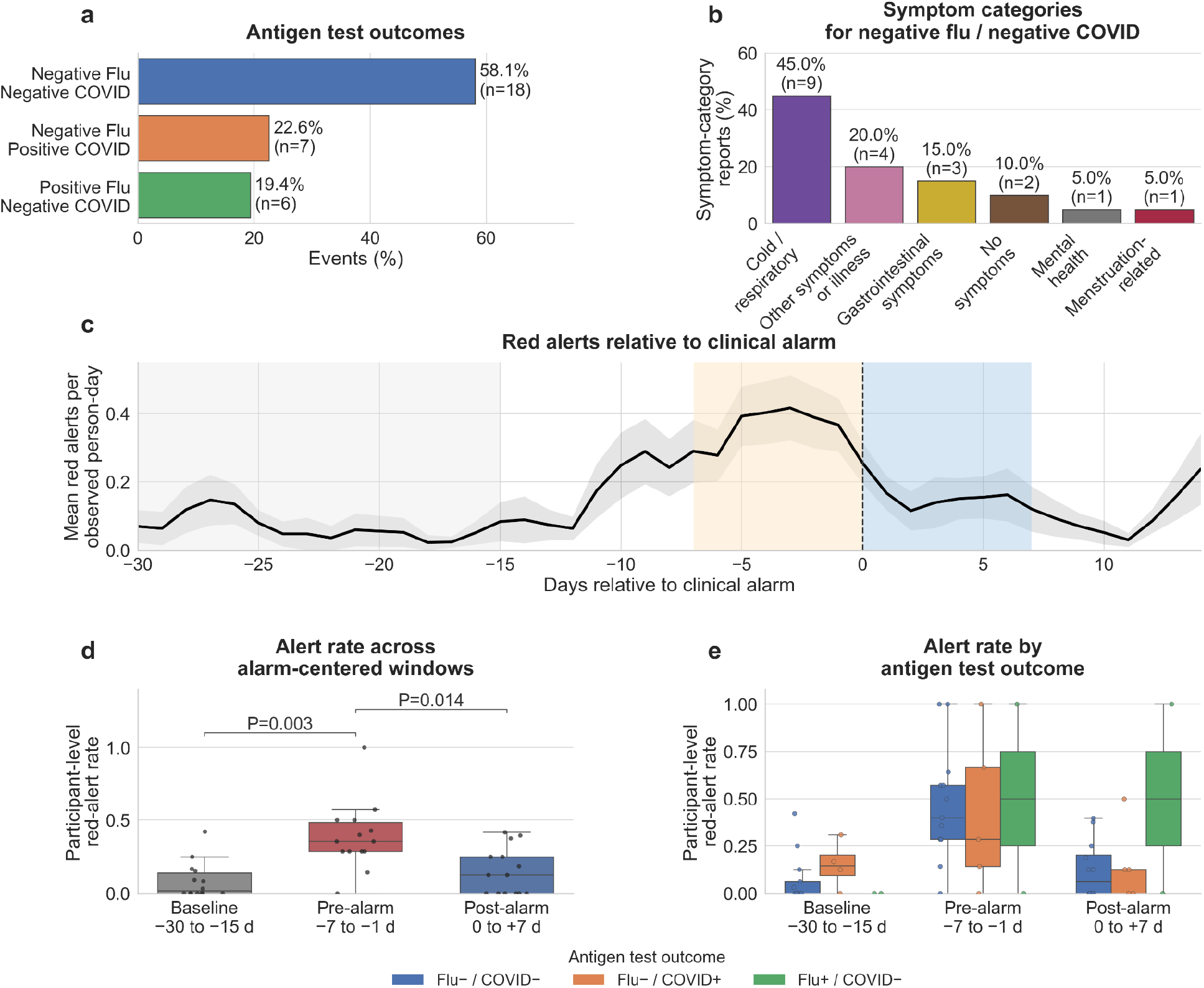
Outcomes of the alerting algorithm and antigen testing. **a**, Distribution of 31 antigen-test outcomes among participants with reported antigen test outcomes. Bars show the percentage of outcome records in each antigen-test outcome category, with counts shown next to each bar. **b**, Symptom categories reported among participants with negative influenza and negative SARS-CoV-2 antigen-test results; 20 symptom-category reports were recorded across the 18 antigen-negative events. Bars show the percentage of reports in each symptom category, with counts shown above each bar. **c**, Participant-weighted daily red alert trajectory aligned to the clinical alarm date. The black line shows the mean of participant-level centered 3-day smoothed rates, and the shaded band indicates ± 1 standard error across those values, with the lower bound clipped at zero. Days are displayed when at least five values (smoothed by participant) contribute. The vertical dashed line denotes the clinical alarm date, and background shading indicates the baseline window (days −30 to −15; gray), pre-alarm window (days −7 to −1; orange), and post-alarm window (days 0 to 7; blue). **d**, Participant-level red alert rates across the three alarm-centered windows. The analysis included 14 participants contributing 21 events with observed NightSignal data in all three windows. Rates from repeated events were averaged within participant so that each point represents one participant. Boxes show the median and interquartile range. Brackets show significant pairwise Wilcoxon signed-rank comparisons after Holm adjustment. **e**, Participant-level red alert rates across windows, stratified by antigen test outcome. Boxes show the median and interquartile range, and points represent participant-level values. In the legend, Flu and COVID denote influenza and SARS-CoV-2 antigen results, and −/+ denote negative/positive results.

We examined NightSignal red alerts aligned to the recorded date on which the clinical alarm was received by the study team (see Methods). A red alert denotes a daily NightSignal algorithm output, and a clinical alarm denotes the alarm recorded as received by the study team. Here, an event denotes an included antigen-test outcome record, and day 0 is the recorded clinical alarm date. The baseline, pre-alarm, and post-alarm windows comprise days −30 to −15, −7 to −1, and 0 to +7, respectively. The available data included 25 events from 17 participants. Because clinical alarms were based on consecutive red alerts, the pre-alarm rise is expected and partly reflects event selection and the operational trigger. The highest smoothed red alert rate occurred 3 days before the clinical alarm (0.417 red alerts per observed participant-day; SE = 0.095; 16 contributing participant-smoothed values; **Figure 2c**). Window comparisons included 21 events from 14 participants, with each included event having at least one NightSignal day in each of the three analysis windows (**Figure 2d**). Median participant-level red-alert rates were lowest during the baseline window from days −30 to −15 (median = 0.016, IQR = [0.000, 0.139]), increased during the pre-alarm window from days −7 to −1 (median = 0.357, IQR = [0.286, 0.482]), and decreased (but not to baseline levels) after the alarm date during days 0 to 7 (median = 0.125, IQR = [0.000, 0.250]; **Figure 2d**). The overall difference in alert burden across these three time windows was statistically significant (Friedman test, P = 0.000843). Pairwise comparisons further showed that the pre-alarm window had significantly higher alert rates than the baseline window (Holm-adjusted P = 0.00256) and the post-alarm window (Holm-adjusted P = 0.0143). Alert rates in the post-alarm window did not differ significantly from those in the baseline window (Holm-adjusted P = 0.185).

When stratified by antigen test outcomes, median participant-level red-alert rates in the antigen-negative group increased from 0.000 (IQR = [0.000, 0.063]; n = 13 participants) during baseline to 0.400 (IQR = [0.286, 0.571]; n = 13) during the pre-alarm window, before decreasing to 0.063 (IQR = [0.000, 0.203]; n = 12) in the post-alarm window (**Figure 2e**). In the COVID-positive group, median rates were 0.146 (IQR = [0.094, 0.202]; n = 4) during baseline, 0.286 (IQR = [0.143, 0.667]; n = 5) during the pre-alarm window, and 0.125 (IQR = [0.000, 0.125]; n = 5) during the post-alarm window. In the influenza-positive/COVID-19-negative group, median rates were 0.000 (IQR = [0.000, 0.000]) during baseline, 0.500 (IQR = [0.250, 0.750]) during the pre-alarm window, and 0.500 (IQR = [0.250, 0.750]) during the post-alarm window, although only two participants contributed to each window. Exploratory participant-clustered comparisons identified an overall difference among antigen-test outcome groups during baseline (Wald χ^2^(2) = 12.74, Holm-adjusted P = 0.005), but not during the pre-alarm (Wald χ^2^(2) = 0.07, Holm-adjusted P = 1.000) or post-alarm window (Wald χ^2^(2) = 1.13, Holm-adjusted P = 1.000). No individual pairwise comparison remained significant after Holm adjustment across all nine outcome-group contrasts (minimum adjusted P = 0.057). The apparent baseline difference should be interpreted cautiously because the influenza-positive group included only two participants, both with zero baseline red-alert rates.

### Activity-adjusted heart rate rises before the clinical alarm

We examined alarm-aligned trajectories of wearable HR features across the window spanning 30 days before to 14 days after the clinical alarm date (**Figure 3**). Four features relating to resting or average HR were analyzed (see Methods): median HR, minimum RHR, 5th percentile HR, and the HR intercept from the HR-activity association model (representing estimated HR at zero steps). Aligned data were available for 15 participants for median HR, 11 participants for minimum RHR, 17 participants for 5th percentile HR, and 16 participants for estimated HR at zero steps. Smoothed daily trajectories showed increases in several HR features in the days preceding the alarm date (**Figure 3a-d**). Within the pre-alarm window, the highest smoothed baseline-relative mean for estimated HR at zero steps occurred 4 days before the alarm (+2.63 bpm), while minimum RHR and 5th percentile HR reached their highest values 3 days before the alarm (+2.09 bpm and +1.63 bpm, respectively), and median HR reached its highest value 4 days before the alarm (+1.53 bpm). These patterns are consistent with an elevation in basal and activity-adjusted HR during the pre-alarm window.

**Figure 3.**
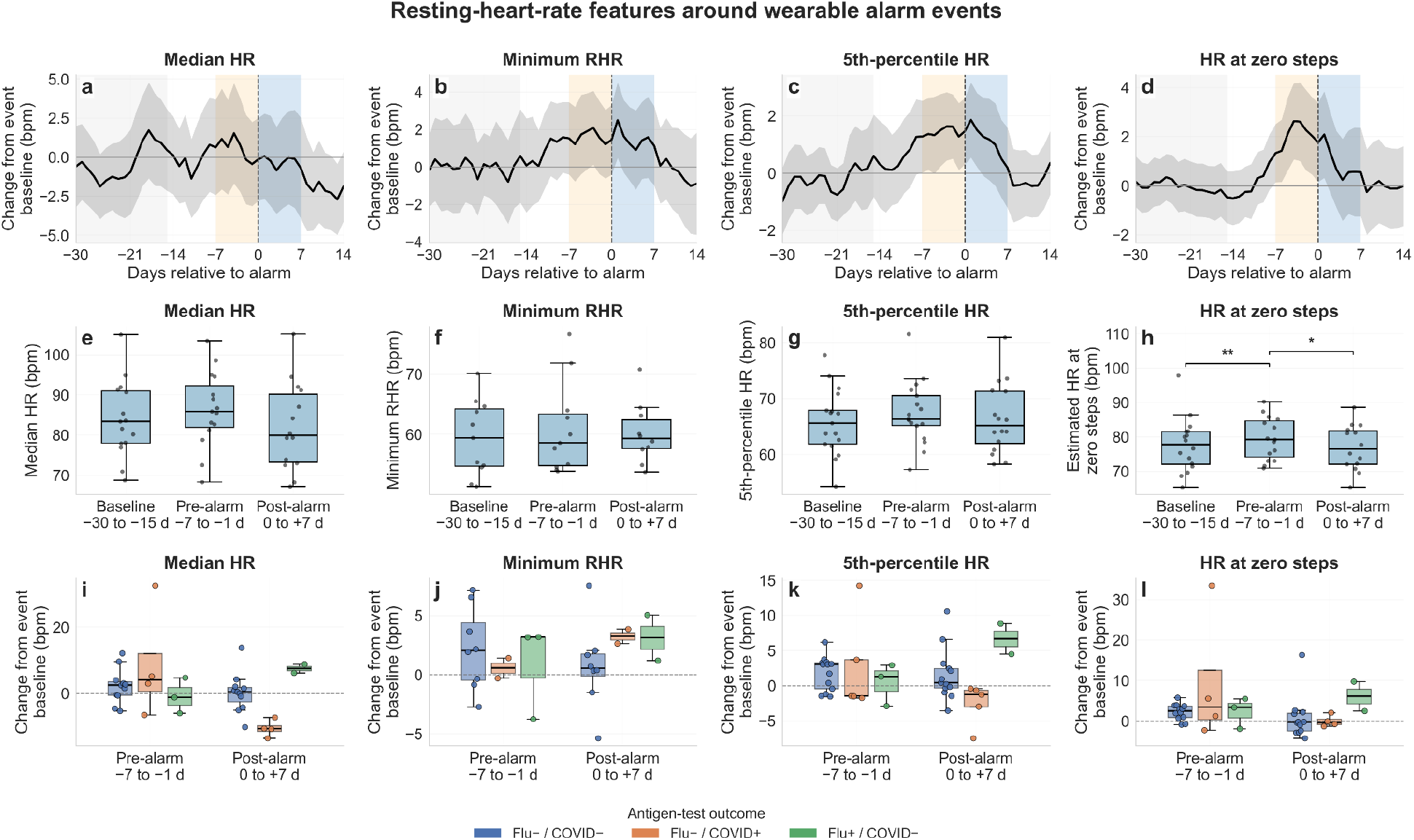
Resting heart rate features around clinical alarm events. **a-d**, Centered 7-day rolling mean changes in daily HR features from each event’s baseline mean (days −30 to −15), aligned to the clinical alarm date (day 0). The black line shows the centered 7-day rolling average of daily participant-mean changes from event baseline in median HR (**a**), minimum RHR (**b**), 5th percentile HR (**c**), and estimated HR at zero steps (**d**). Gray shaded bands indicate standard error, and the vertical dashed line marks the date of clinical alarm (day 0). Gray, orange, and blue backgrounds indicate baseline, pre-alarm, and post-alarm windows. **e-h**, Comparisons of absolute participant-level HR feature window means relative to the alarm date. Boxes show the median and interquartile range, data points represent individual participant values. Brackets indicate paired Wilcoxon signed-rank comparisons after a significant Friedman test, with Holm adjustment across the three comparisons within each feature (* P < 0.05, ** P < 0.01, *** P < 0.001). **i-l**, Participant-level changes from event baseline during the pre-alarm and post-alarm windows, stratified by antigen-test outcome. In the legend, Flu and COVID denote influenza and SARS-CoV-2 antigen results, and −/+ denote negative/positive results.

Window-based analyses also supported a pre-alarm increase in certain HR features (**Figure 3e-h**). Estimated HR at zero steps increased from 77.7 (SD = 7.9) bpm during the baseline window to 79.5 (SD = 6.2) bpm during the pre-alarm window, then decreased to a baseline-like value of 76.9 (SD = 6.6) bpm in the post-alarm window. The increase from baseline to pre-alarm window was significant (Holm-adjusted Wilcoxon signed-rank P = 0.00787, median paired change = 2.48 bpm), as was the decrease from the pre-alarm to post-alarm window (Holm-adjusted P = 0.0405, median paired change = −3.02 bpm). The 5th percentile HR showed a similar modest pre-alarm increase, rising from 65.7 (SD = 5.9) bpm at baseline to 67.4 (SD = 5.6) bpm in the pre-alarm window, before decreasing to 66.4 (SD = 6.3) bpm post-alarm. The paired increase from baseline to pre-alarm did not remain significant after Holm adjustment (P = 0.080, median paired change = 1.72 bpm). Median HR and minimum RHR showed directionally similar pre-alarm increases but did not reach statistical significance in the overall paired window comparisons. Specifically, median HR increased from 84.1 (SD = 9.5) bpm at baseline to 86.4 (SD = 9.4) bpm in the pre-alarm window, while minimum RHR increased from 59.3 (SD = 6.2) bpm to 60.8 (SD = 7.5) bpm. When stratified by antigen test outcome, the clearest changes occurred in the antigen-negative group, which had the largest sample size (**Figure 3i-l**). In the antigen-negative subset, estimated HR at zero steps differed across windows (Friedman p = 0.0388), with an increase from baseline to the pre-alarm window (median paired change = 2.52 bpm, Holm-adjusted P = 0.0146). The baseline to pre-alarm increase in 5th percentile HR did not remain significant after adjustment (median paired change = 3.03 bpm, adjusted P = 0.0798). These analyses stratified by test outcome were considered exploratory given the small subgroup sizes.

### HR/activity relationship and additional HR features remain largely stable around the alarm

We examined the coupling of HR and activity features around the clinical alarm date. Across the full aligned window, daily steps were positively associated with the variability in HR across the day (HR SD; beta = 0.642, SE = 0.046, Holm-adjusted P < 0.001) and 95^th^ percentile of HR (beta = 0.659, SE = 0.046, Holm-adjusted P < 0.001) (**Figure 4a**). These associations were also present in each analysis time window, consisting of the baseline window (HR SD: beta = 0.658, SE = 0.069, Holm-adjusted P < 0.001; 95th percentile HR: beta = 0.640, SE = 0.070, Holm-adjusted P < 0.001), the pre-alarm window (HR SD: beta = 0.576, SE = 0.107, Holm-adjusted P < 0.001; 95th percentile HR: beta = 0.554, SE = 0.113, Holm-adjusted P < 0.001), and the post-alarm window (HR SD: beta = 0.634, SE = 0.098, Holm-adjusted P < 0.001; 95th percentile HR: beta = 0.646, SE = 0.091, Holm-adjusted P < 0.001). Thus, higher variability in HR and higher upper-tail HR throughout the day were consistently associated with higher daily activity throughout the time windows around the alert. The within-day HR-steps correlation was also positively associated with daily steps when examined across the full aligned window (beta = 0.246, SE = 0.057, Holm-adjusted P < 0.001) and during the baseline window (beta = 0.284, SE = 0.080, Holm-adjusted P = 0.0027). However, this association was not statistically significant when analyzed within the pre-alarm window or post-alarm window after Holm adjustment. Nocturnal HR slope from 8pm to minimum HR was also not significantly associated with daily steps, either overall or within the specific windows (all Holm-adjusted P > 0.05, **Figure 4a**).

**Figure 4.**
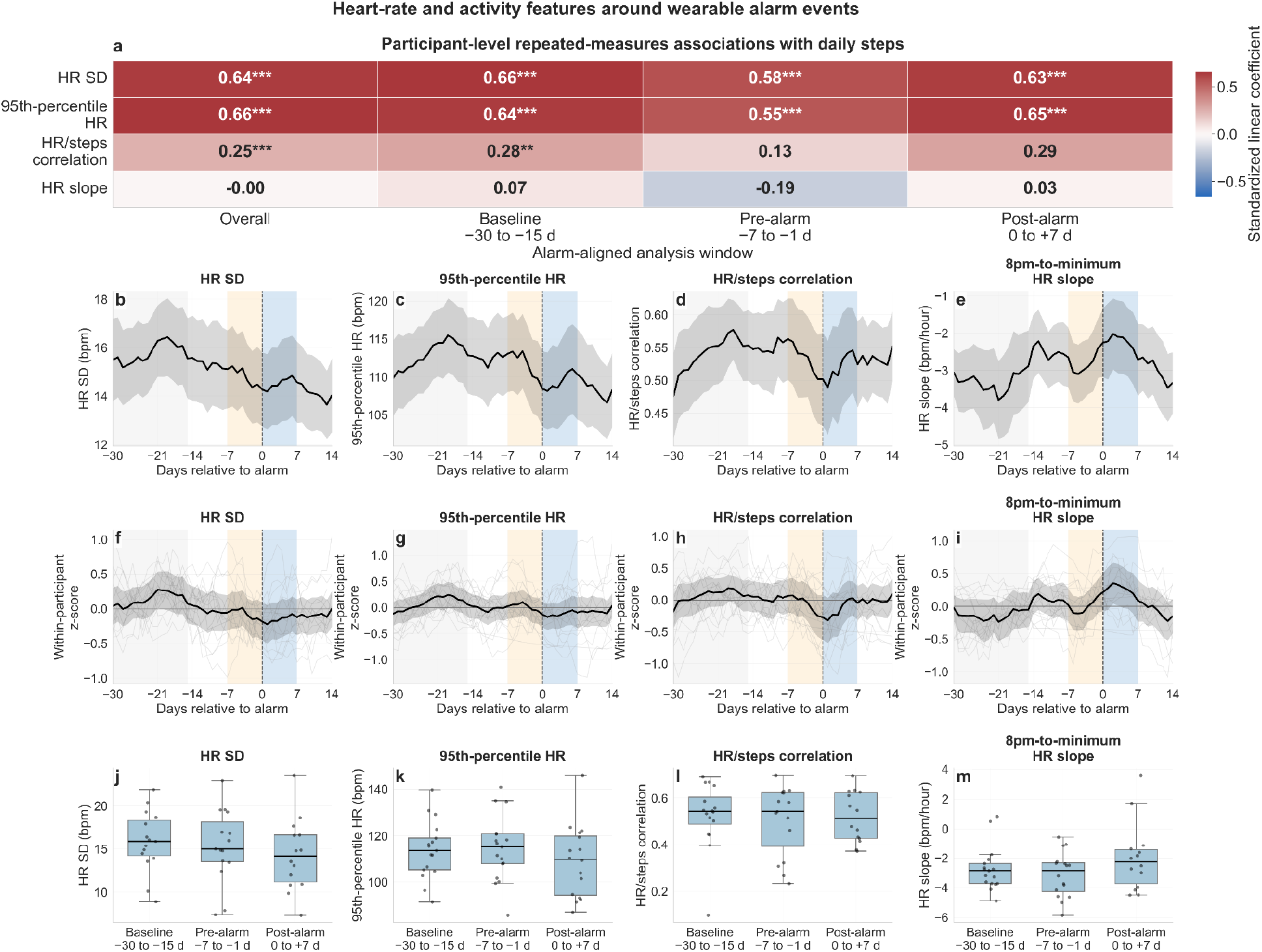
Further HR features and HR/activity relationship. **a**, Linear mixed-effects models were used to examine daily associations between selected wearable HR features and daily steps after alignment to the clinical alarm date. Heatmap values show standardized regression coefficients, and asterisks denote significance (based on Holm-adjusted P-values; * denotes P < 0.05, ** denotes P < 0.01, *** denotes P < 0.001). Associations are shown overall (first column) and within analysis windows (baseline, pre-alarm, and post-alarm). **b-e**, Mean trajectories in the original feature units of selected wearable features from 30 days before to 14 days after clinical alarm date. Repeated events were averaged within participant before participant-level summaries. Lines show the rolling average for all participants with an alert and associated test, and shaded regions indicate standard error of the mean. **f-i**, Dimensionless within-participant z-score trajectories for the same features, showing deviations from each participant’s typical level. Gray lines show individual participant trajectories after smoothing, black lines show the all-participant rolling average, and shaded regions denote the standard error of the mean. In **b-i**, the dashed line marks the clinical alarm date; gray, orange, and blue backgrounds indicate baseline, pre-alarm, and post-alarm windows. **j-m**, Feature summaries across the pre-detection baseline, pre-alarm, and post-alarm windows. Each point represents a participant-level window average, and boxes summarize the distribution across participants (median and interquartile range). Overall window differences were assessed using Friedman tests followed by Holm-adjusted Wilcoxon signed-rank tests.

We assessed temporal trajectories and participant-level summaries of the selected wearable features across the period surrounding the alert and infection. Rolling-average trajectories suggested modest variation across the alert-aligned time course, with decreases in HR SD and 95th percentile HR around and after the alarm date (**Figure 4b** and **Figure 4c**), a decrease in Pearson R (from the HR and steps correlation) around the alarm date with a recovery after (**Figure 4d**), and an increase in the 8pm to minimum HR slope around or slightly after alarm (**Figure 4e**). The same patterns, along with individual trajectories, were observed in the smoothed z-scores (**Figure 4f-i**). The paired participant-level comparisons across the baseline, pre-alarm, and post-alarm windows were not significant for any of the four features (Friedman P > 0.05 for all; **Figure 4j-m**), and no pairwise comparison remained significant after Holm adjustment. Nocturnal 8pm-to-minimum HR slope was higher post-alarm than pre-alarm in the unadjusted pairwise comparison (Wilcoxon P = 0.035), although this association did not remain significant after Holm adjustment (adjusted P = 0.106).

### Sleep and circadian rhythm features are disrupted around the alarm

Wearable-derived circadian features showed evidence of modest perturbation around the clinical alarm events. Across all alarm events, mesor (the midline in the circadian fit; see Methods) differed across the baseline, pre-alarm, and post-alarm windows (Friedman P = 0.028), although the baseline-to-pre-alarm increase did not remain significant after Holm adjustment (median paired change = 3.40 bpm; unadjusted P = 0.035, adjusted P = 0.104; **Figure 5c**). Within the antigen-negative group, mesor also differed across the windows (Friedman P = 0.037), with a significant increase from baseline to pre-alarm (median paired change = 3.26 bpm, Holm-adjusted P = 0.010). The goodness of fit of the daily cosinor model, R^2^, was most altered after the alarm date. Across all participants, the overall difference in R^2^ was not significant (Friedman P = 0.099). However, within the antigen-negative group, R^2^ differed across windows (Friedman P = 0.0058), with a decrease from baseline to the post-alarm window (median paired change = −0.072, Holm-adjusted P = 0.005; **Figure 5d**). Circadian amplitude and acrophase showed less consistent evidence of change in the pre-alarm or post-alarm windows (**Figure 5a,b**). Amplitude displayed a steady decrease until around or slightly after the alarm date, and then increased (**Figure 5e**), while acrophase was relatively stable and then declined after the alarm date (**Figure 5b,f**). The mesor spiked prior to or around the alarm date and then decreased after the alarm (**Figure 5c,g**), while the goodness of fit R^2^ feature displayed a steady decrease until around the alarm date and then increased (recovered) after (**Figure 5h**). To illustrate sleep disruption at the individual level, one participant with a positive COVID outcome was selected from participants with a positive influenza or SARS-CoV-2 outcome according to availability of both sleep features and the number of observed feature values. This case study showed an increase in WASO/TST from 2.72% during baseline to 10.45% during the pre-alarm window, and 9.82% during the post-alarm window, with an increase in the number of awakenings from 5.64 at baseline to 7.86 pre-alarm and 9.29 post-alarm (**Figure 5i,j**). Together, these results suggest that wearable-derived circadian and sleep features capture heterogeneous circadian/sleep disruption and individualized physiological perturbations around wearable alarm events.

**Figure 5.**
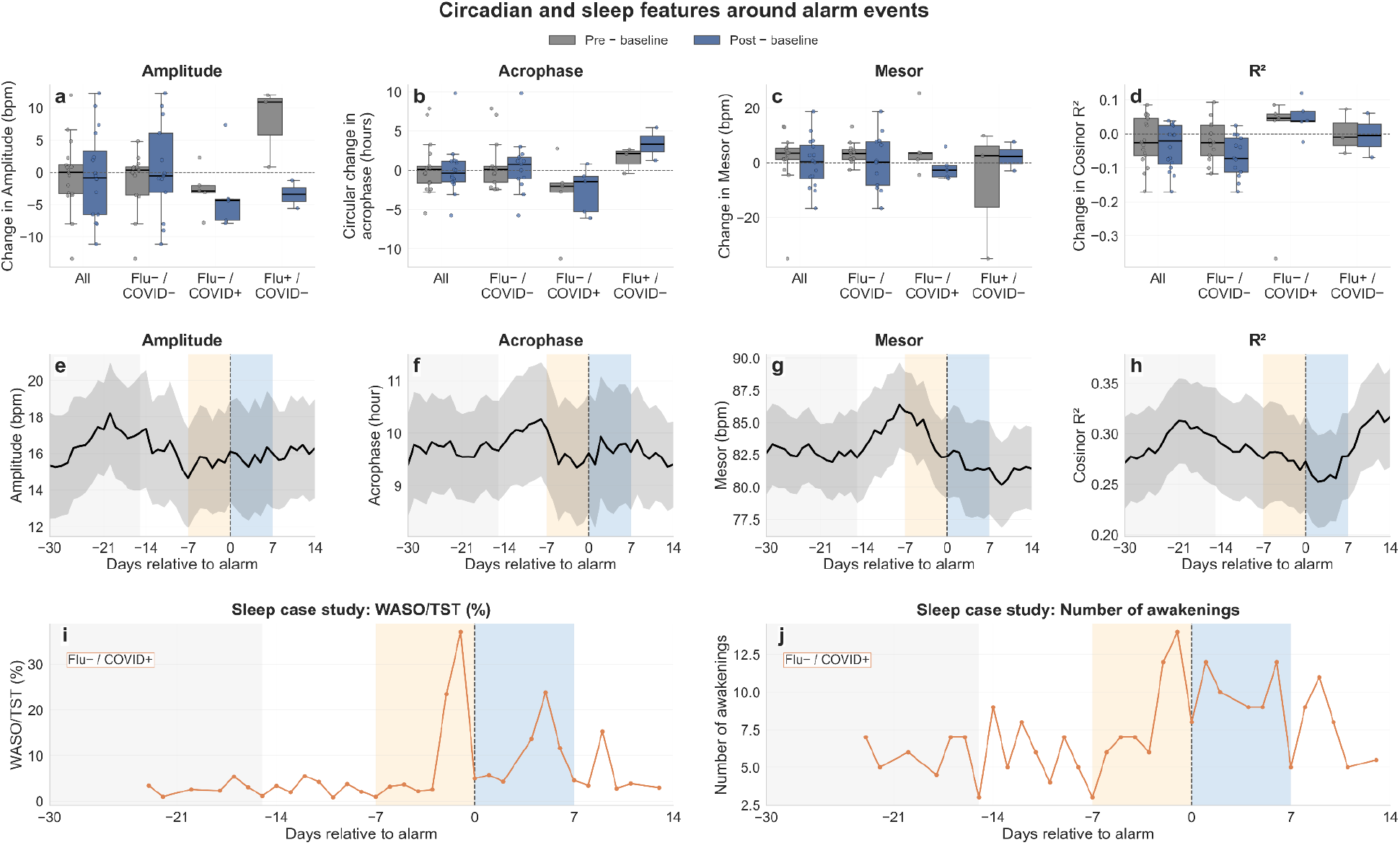
Circadian rhythm and sleep features around alarm events. **a-d**, Participant-level changes in daily cosinor-derived features aligned to the clinical alarm date. Changes are shown for all participants and separately by antigen-test outcome relative to each event’s baseline mean from days −30 to −15. Gray boxes show the change during the pre-alarm window from days −7 to −1, and blue boxes show the change during the post-alarm window from days 0 to 7. Features shown are the cosinor amplitude (**a**), circular acrophase (**b**), mesor (**c**), and model fit (R^2^) (**d**). Boxes show the median and interquartile range, whiskers extend to the most extreme values within 1.5 interquartile ranges of the box, and points represent individual participants. Repeated events were averaged within participant before participant-level summaries. Window comparisons were assessed using Friedman tests followed by paired Wilcoxon signed-rank tests with Holm adjustment, except for acrophase, which used paired Wilcoxon tests of signed circular differences with Holm adjustment. **e-h**, Mean daily trajectories of cosinor features across all participants, aligned to the alarm date. Lines show 7-day centered rolling means, and shaded bands show the corresponding smoothed standard error. **i**,**j**, Sleep feature trajectories for one illustrative SARS-CoV-2-positive participant, showing WASO/TST expressed as a percentage (**i**) and number of awakenings (**j**). Points indicate available participant-level values by relative day, and lines connect available observations. In **e-j**, the vertical dashed line denotes the clinical alarm date; gray, orange, and blue backgrounds indicate the baseline, pre-alarm, and post-alarm windows. Flu and COVID denote influenza and SARS-CoV-2 antigen results, and −/+ denote negative/positive results.

## DISCUSSION

We evaluated the feasibility, deployment, and observed antigen-test outcomes of the NightSignal algorithm, a real-time wearable alerting system, for detecting physiological changes associated with infection among pediatric transplant recipients and their household members. The alerting algorithm was implemented prospectively, alerts were tracked during active monitoring, and participants were contacted and tested in the case of sustained red alerts. While only 13/31 (41.9%) of the red alert events that yielded participant contact and antigen testing corresponded to a confirmed influenza or COVID infection, the remaining 18 antigen-negative events were almost always (16/18, 88.9%) accompanied by respiratory, gastrointestinal, or other reported symptoms. This highlights the ability of the method to capture clinically relevant physiological disturbances in a real-world setting and a vulnerable population. By linking smartwatch-based alerts to at-home antigen testing, wearable features, and clinical study team response, this study evaluates wearable-guided infection surveillance in high-risk pediatric transplant households. The present report focuses on wearable alerting and associated physiology; baloxavir treatment uptake, timing, and efficacy were not evaluated in these analyses.

The algorithm was responsive to systemic physiological stress and illness, but alerts were not specific to a particular pathogen or set of pathogens. Thus, a wearable alert should not be interpreted as evidence specific to influenza or COVID or as a diagnosis of either infection. Instead, it functions as a signal that an individual’s physiology has deviated from its typical (baseline) range, and that actions such as symptom assessment, testing, masking, reducing exposure to vulnerable individuals, or closer clinical monitoring may be warranted. This study did not capture comprehensive contextual data of participant behaviors, mental state, or other confounding factors around the alert periods. Users of the alerting system may therefore be able to better contextualize changes in their wearable physiology with lifestyle events or behaviors (e.g., holidays, stress, schedule changes) that may trigger non-infection alerts. The original NightSignal study similarly found that alerts can precede viral infections but may also correspond to other physical or emotional stressors that alter physiology ^29^. The alerting algorithm was relatively simple and interpretable, relying on individualized deviations in nighttime RHR. In contrast to more complex machine learning models, the present approach can be readily inspected, communicated to users and clinicians, and implemented across devices that provide HR and activity time series data.

A major contribution of this study was the implementation of a system that acquired wearable data, applied a personalized detection algorithm, and generated alerts with limited researcher or participant involvement. This degree of automation is important for both user acceptability and research scalability. In the future, improvements to these systems could further increase automation and decrease the burden on participants, clinical teams, coordinators, and researchers. These improvements could include transmitting the alerts directly to the participant and/or clinician, eliminating the need for participants to open an app to upload data, and automatically detecting data missingness periods. Low-wear and missing data are frequent challenges in longitudinal and real-world wearable studies, due to device removal for charging or showering, loss of charge, device malfunction, or synchronization issues ^30,31^. These concerns are particularly relevant for real-time alerting or anomaly detection studies, where the alarm system cannot function without access to recent wearable data. The need to make the system as easy as possible for the user and clinician may also be heightened for patient populations like this one, where participants may be facing their own medical challenges and have varying levels of comfort with technology, and clinicians may not have the bandwidth to manually monitor wearable data or engage with alerting logistics. Device removal during sleep, potentially due to discomfort, may also have contributed to sparse sleep data.

In addition to alert results, we examined wearable physiology around alarms to gain insight into changes associated with infection and additional signals that could be used to improve performance in next-generation algorithms. The observed elevations in RHR-related features around alerts and infection events were consistent with the expected autonomic and inflammatory response to acute illness. RHR may increase as a consequence of fever, immune activation, stress, or other factors ^32–34^. Indeed, the overall pattern of a spike in RHR features at or around the clinical alarm or symptom date (followed by a decrease thereafter) matches similar patterns shown in prior work ^20,21,35^. Additionally, joint analysis of HR and activity data may help clarify changes in their relationship around infection. Prior work has incorporated an HR over steps measure ^20^ or examined HR increase per step ^21^ during COVID infection. In particular, these features may capture an alteration in the usual linkage between cardiovascular demand and movement. During infection, individuals may exhibit a greater HR response for a given amount of activity, consistent with reduced cardiopulmonary efficiency, systemic physiological stress, or incomplete recovery ^20,21,36^. Here, we did not observe similarly clear changes in the HR/activity coupling: while the HR/steps correlation coefficient appeared to decrease around the alarm date, values across the analysis windows were not significantly different. The decline in HR during the evening/night appeared steeper during the pre-alarm window than during the post-alarm window, although this difference did not remain significant after adjustment for multiple comparisons (**Figure 4m**).

We identified changes in cosinor-derived HR features around the alarm date and described sleep features in one illustrative participant. The circadian goodness of fit measure showed a post-alarm decrease that was most evident in the antigen-negative group (although the overall participant-level comparison across windows was not significant). This lower R^2^ indicates poorer fit of the 24-hour harmonic model to the observed HR data during these peri-alert periods, i.e., circadian or diurnal rhythmicity in HR may have been less clearly expressed. This could be due to early effects of infection (e.g., immune activation, behavioral changes) disrupting the typical circadian rhythm ^37–39^. The amplitude trajectory also appeared to decrease around the alarm date, likewise potentially reflecting a weakened HR rhythm. The mesor was higher in the pre-alarm window than at baseline, although this comparison did not remain significant after Holm adjustment; as the mesor reflects the average HR after accounting for 24-hour variations, this pattern could similarly reflect RHR-related changes due to immune response, fever, and stress ^32–34^. The illustrative COVID-positive participant showed marked sleep disruption across the peri-alarm period, highlighting the potential for substantial individual-level sleep quality changes.

These results support expanding future versions of the algorithm beyond or alongside nighttime RHR. In addition to these HR, activity, and sleep/circadian features, modern wearable devices frequently capture HRV, respiratory rate, oxygen saturation, skin temperature, and other signals. HRV may be particularly informative because it reflects autonomic regulation and may decrease around infection ^40–42^; future studies are needed to collect HRV and infection data in larger cohorts. Respiratory rate, oxygen saturation, and temperature provide additional signals directly related to respiratory infection ^43–45^, although accuracy and availability differ across wearable devices.

This study has several limitations. Because clinical alarms were linked to sustained elevations in nighttime RHR, pre-alarm increases in red-alert burden and related HR features are partly expected from event selection, and subsequent decreases may reflect regression to the mean. Participant-level event averaging and random intercepts do not account for dependence between household members. The number of antigen-confirmed influenza and COVID infections was small, limiting pathogen-specific inference and the precision of performance estimates. The cohort size and focus on pediatric transplant households may hinder generalizability, and larger studies are needed. Additionally, antigen-negative symptomatic events could have represented other respiratory pathogens, false-negative antigen tests, or other physiological stressors. More comprehensive laboratory testing would be needed to determine the root cause of these events. Immunosuppressive regimens were not available for analysis, precluding assessment of their association with wearable features or alert burden. Wearable adherence and data synchronization displayed substantial interindividual variability; this is a major concern, since alerts could only be generated on days when current wearable data were present. As this was a feasibility trial and individuals were not regularly tested outside of the alerts or the development of symptoms, sensitivity and specificity could not be established. A larger study with systematic reference testing could assess false-negative and false-positive alerts and diagnostic performance. Device heterogeneity may impact sampling density and the comparability of derived features. Finally, RHR and related wearable signals can be impacted by a variety of confounding variables (e.g., exercise, postural changes, sleep loss, psychological stress, medication, vaccination) ^29,46–50^. Future studies that collect more detailed logs of the possible confounding factors on a daily or weekly basis may be needed to better distinguish true infection-related wearable patterns.

Overall, this study demonstrates that automated, real-time wearable alerting is feasible in transplant recipients and their household members and identifies physiological changes around clinical alarm events. The occurrence of confirmed influenza, COVID, and other symptom reports among alerted participants suggests that the system detected meaningful departures from usual physiology. The observed changes in HR, its relationship with activity, and sleep/circadian features further indicate that the response to infection is multidimensional. Incorporating activity-adjusted cardiovascular measures, circadian rhythms, and other physiological signals such as HRV may improve pathogen-specific detection while preserving the interpretability, scalability, and low participant burden of the current system. This wearable alerting system could serve as an additional layer of surveillance that supports timely testing, precautionary behavior, and clinical monitoring in populations at elevated risk from infection.

## Data availability

Summaries of the extracted smartwatch features necessary to reproduce the manuscript analyses and figures will be available at https://github.com/mayercl/infection-transplant upon publication. Additional data produced in the present study are available from the corresponding author on reasonable request.

## Code availability

The underlying code for this study is planned for release at https://github.com/mayercl/infection-transplant upon publication, and archived in a DOI-minting repository. The analysis was performed in Python (v.3.11.4; Jupyter Notebook) using packages including numpy, pandas, matplotlib, seaborn, scipy, scikit-learn, statsmodels, and CosinorPy.

## Acknowledgements

This Phase IV clinical trial was funded by Roche-Genentech. This parent study was supported by UnitedHealthGroup. C.Ma. is grateful for support by the NIH institutional research training grants (2T32AR007422-41 and DK007217-49). We thank the participating families and the clinical study staff.

## Author contributions

M.P.S. and B.K. conceived the study. M.P.S., S.A., M.J.O., and B.K. developed the clinical protocol and oversaw participant recruitment, consent, and clinical follow-up. Ek.S.R. contributed to early study planning. C.B., M.A.-M., A.C., and B.K. coordinated wearable device distribution, participant support, antigen testing, and clinical data collection. H.G. facilitated access to the REDCap clinical data. C.Ma. processed wearable data and alerting information. C.Ma., E.H., M.P.S., and B.K. led the wearable data analysis and developed the analytical methodology. T.B. and Es.R. contributed to wearable planning and analysis. C.Ma. generated the visualizations and wrote the original manuscript draft. C.Ma., E.H., T.B., Es.R., Ek.S.R., H.A., K.-M.V., V.M., A.A.S., C.My., S.A., M.J.O., M.P.S., and B.K. contributed to manuscript review and revision. All authors reviewed and approved the final manuscript.

## METHODS

### Study design and participants

The Phase IV clinical trial for Xofluza (baloxavir marboxil) “A Pilot Study of Xofluza treatment of Influenza in Pediatric Transplant Recipients and Household Members after early infection alerting using Wearable Devices” was approved by the Children’s Hospital of Philadelphia Institutional Review Board (IRB#22-020440). This Phase IV clinical trial was funded by Roche-Genentech. IRB#22-020440 is a sub-study of an ongoing study “Early Detection of SARS-CoV-2 & other Infections using Wearable Devices in Pediatric Transplant Patients and Household Members” (IRB 20-017872, PI: Brendan Keating). This parent study was supported by UnitedHealthGroup.

IRB#22-020440 was a prospective, single-center feasibility study of wearable-based early infection detection, alerting, rapid home respiratory virus testing, and clinically guided antiviral treatment. The cohort consisted of pediatric solid-organ transplant recipients, transplant waitlisted patients, and their household members. The household members were included to capture both high-risk transplant recipients and household contacts who may contribute to respiratory virus transmission or exposure. Participants were recruited from the clinic population, with potentially eligible transplant recipients and waitlisted patients identified through clinical screening by the study coordinator and clinical team. Eligible patients and their household members were enrolled after informed consent, with separate consent forms distributed to each participating household member. The study population included kidney, heart, liver, and lung transplant recipients. Participants were provided wearable devices (see *Wearable devices and wearable data* section below) for continuous physiological monitoring, and provided at-home respiratory virus antigen tests. Following enrollment, the study team documented household enrollment information, participant identifiers, device assignment, shipment details, date offset values, and MyPHD keys in REDCap (see more detail about MyPHD below). Study staff confirmed receipt of devices and test kits, distributed device setup instructions and MyPHD keys, and contacted families to confirm setup. Participants were tracked virtually during the study period via wearable infection alerts (NightSignal algorithm^29^), symptoms, antigen test outcomes, antiviral use, and clinical outcomes. If wearable alerting suggested possible infection, the study team documented the alert and subsequent actions in REDCap, including the date of the alarm, alarm signal report, action taken, antigen test outcome, date of antigen testing, and additional notes. When participants tested positive for influenza, the study team contacted the participant and/or family to determine whether the participant wished to take baloxavir marboxil (Xofluza) and coordinated with the study physician, transplant clinical team, and participant’s primary care physician as appropriate. Participants who received Xofluza continued to be monitored according to study procedures.

### Wearable devices and wearable data collection

Participants were provided consumer-grade wearable devices, including Apple Watch and Fitbit devices, for longitudinal physiological monitoring. Devices and respiratory virus test kits were packaged and shipped to participant family members. Participants received email instructions for device setup, including individualized MyPHD keys, and study coordinators contacted families to confirm that devices were successfully connected and transmitting data.

Wearable data were collected through the My Personal Health Dashboard (MyPHD) infrastructure, which enabled ingestion and processing of smartwatch biometric data ^51^. The primary signal used for infection alerting consisted of HR, particularly RHR patterns during nighttime. Additional signals from the devices included steps/activity, sleep, HRV, proxies of body temperature, oxygen saturation, respiration, and other device-dependent data types. Wearable monitoring was intended to occur continuously during the study period. Participants were sent automated daily reminders through MyPHD to upload/sync their wearable data. Wearable data were linked to study records using participant-specific identifiers and MyPHD key files. Study staff maintained study management information in REDCap, including the participant identifier, household enrollment information, device assignment, and date offset values needed for downstream wearable data processing. Alerts were sent to the study team on a daily basis, and if three or more consecutive red alerts were received for a participant, the study team contacted the participant or household to ask whether the participant was experiencing flu-like symptoms. Symptoms, testing actions, antigen test results, and follow-up notes were documented in REDCap.

### Alerting algorithm

The NightSignal algorithm was used for the real-time wearable infection alerting ^29^. This previously published algorithm uses within-person deviations in nighttime RHR, to identify periods in which a participant’s biometric data are abnormal relative to their own baseline. A deterministic finite-state-machine framework was used to govern transitions between alert states (red, yellow, green) based on sustained differences in median nighttime RHR. The algorithm was implemented through the MyPHD platform, and alerts were transmitted along with the other wearable data types to a secure Google Cloud Bucket. Alerts were then processed on a daily basis, and recent red alerts were transmitted to the clinical study team. Since recent data were needed to run the NightSignal algorithm through MyPHD, the participant needed to upload/sync their data to MyPHD before the automated run of the algorithm and the alert generation was possible.

### Antigen testing

Participants were provided at-home respiratory virus antigen tests at enrollment. Study shipments included two COVID-19 tests and two influenza tests per family member. When a wearable alert met criteria for participant (patient or household member) contact, the study team instructed the participant or household to perform at-home antigen testing according to the manufacturer instructions. The date of the antigen test administration, antigen test outcome, and any study team actions were documented in REDCap.

### Antigen test outcomes and red alert burden

Antigen test results were summarized as counts and percentages of available outcome records. Symptom category counts and percentages used the number of category reports as their denominator, and categories could overlap within a record. For events with negative influenza and negative COVID testing, symptom categories were defined based on a researcher-defined assessment of relevant clinical team notes corresponding to the event, resulting in the categories of no symptoms, mental health symptoms, menstruation-related symptoms, cold or respiratory symptoms, gastrointestinal symptoms, and other symptoms or illness. An event could contribute to more than one symptom category. These categories were descriptive rather than diagnostic; primary analyses therefore retained grouping by antigen-test outcome. Daily alert data were merged with antigen test outcomes and aligned relative to each event’s clinical alarm date, defined as the recorded date on which the clinical alarm was received by the study team. Analyses were restricted to a window from 30 days before to 14 days after the alarm date. For each event and relative day, an observed NightSignal state was coded as 1 if red and 0 if non-red; days without an observed NightSignal state were treated as missing rather than assigned zero alerts. Daily rates were averaged across repeated events within participant and summarized across participants using a centered 3-day mean. The analysis used the three alarm-centered windows of baseline (days −30 to −15), pre-alarm (days −7 to −1), and post-alarm (days 0 to 7). Days −14 to −8 were omitted from window comparisons but retained in trajectories, and day 0 belonged only to the post-alarm window. Overall differences across the three windows were assessed using Friedman repeated-measures tests among events with at least one observed NightSignal day in each window. Event-level window rates from repeated events were averaged within participant before participant-level inference. Pairwise within-participant comparisons between windows were performed using two-sided Wilcoxon signed-rank tests with Holm adjustment. To evaluate whether red-alert rates differed by antigen test outcomes, exploratory between-group comparisons within each window were performed using Gaussian generalized estimating equations with robust standard errors and participant-level clustering, allowing participants to contribute events to more than one antigen test outcome category. Omnibus Wald test p-values were Holm-adjusted across the three windows, and pairwise contrast p-values were Holm-adjusted across all nine outcome-group comparisons.

### Cohort comparisons

To compare patients with household members, we analyzed the number of days with HR data and each participant’s mean daily minimum resting HR (**Figure 1c-d**). Cohort comparisons used ordinary least squares regression with participant group as the predictor and household-clustered robust standard errors, finite sample covariance and degrees-of-freedom corrections, and t-based inference. Days with HR data were log-transformed, and exponentiated coefficients and confidence intervals give the household-member/patient geometric-mean ratio (146 participants, 78 households). Mean daily minimum RHR was analyzed without transformation, with the contrast reported as patient minus household member.

### Resting heart rate wearable feature trajectories and window-level comparisons

Continuous HR data were processed separately for each participant, to generate (typically day-level) summary features. HR files were identified for each participant, and all available files were concatenated into a single participant-level dataset. Duplicate records were removed before feature extraction. HR observations were grouped by calendar day, and for each day with at least one valid HR measurement, the daily median HR was calculated as the median of all available intraday HR values. The 5th percentile HR was calculated using the distribution of that day’s available HR measurements. For RHR feature extraction, HR measurements were aligned with step data using their recorded timestamps. An HR observation was classified as resting when the sum of recorded steps during the preceding 10 minutes was zero. HR observations without activity during this interval were retained as RHR measurements and grouped by participant and calendar day. Daily minimum RHR was defined as the minimum of the available RHR values on that day. As an alternative to estimate HR independent of activity, a daily estimated HR at zero steps feature was calculated. HR and steps were aggregated into 30-minute intervals, and mean HR and total steps (the sum of step counts) were computed within each interval. Only intervals containing at least one HR measurement and at least one step count measurement were retained. For each participant-day with at least six valid intervals and variation in both interval-level HR and steps, a linear regression model was fit with mean HR as the dependent variable and total steps as the independent variable. The model intercept was included in downstream analyses as the estimated HR at zero steps.

These daily wearable-derived features (Median HR, Minimum RHR, 5th percentile HR, and estimated HR at zero steps) were aligned to each participant’s clinical alarm event date. For trajectory analyses, daily values were expressed relative to the event-specific mean during the baseline window, and repeated events were averaged within participants before calculating participant-level means and standard errors. Feature trajectories were summarized across participants for each day relative to the alert, with daily means and standard errors smoothed using a centered 7-day rolling average. For window-based analyses, the windows of baseline (days −30 to −15), pre-alarm (days −7 to −1), and post-alarm (days 0 to 7) were used. Each feature was averaged within each event and analysis window, and repeated event-level values were then averaged within participant. Overall differences across the three windows were evaluated using Friedman tests, followed by two-sided paired Wilcoxon signed-rank tests with Holm adjustment for the three pairwise comparisons within each feature. Analyses were also stratified by antigen-test outcome. Within each outcome group, overall differences across windows were evaluated using Friedman tests, followed by paired Wilcoxon signed-rank tests with Holm adjustment; these outcome-stratified analyses were considered exploratory because of the small subgroup sizes. Formal between-outcome comparisons were not performed because some participants and households contributed to more than one outcome group.

### Heart rate-activity association and trajectory analysis

Daily step counts and daily HR SD features were computed from steps and HR files, respectively, for each participant. All available files for the participant were concatenated, and duplicate records were removed. Daily step count was calculated as the sum of all available intraday step values for the given calendar day, with missing values excluded. Daily HR SD was calculated as the standard deviation of all available intraday HR measurements on the given calendar day. The 95th percentile HR was calculated using the distribution of that day’s available HR measurements. The daily HR and steps correlation feature was calculated using temporally aligned HR and steps. HR and steps were aggregated into 30-minute intervals, and mean HR and total steps (the sum of step counts) were computed within each interval. Only intervals containing at least one HR measurement and at least one step count measurement were retained. For each participant-day with at least six valid intervals and variation in both HR and step counts, Pearson’s correlation coefficient was calculated between interval-level mean HR and total steps. A nocturnal HR slope feature was calculated for each participant and night using HR measurements collected between 8pm and 5am, with measurements after midnight assigned to the preceding night. Within this interval, a centered 10-minute rolling median was applied to the HR measurements, requiring at least three observations within the rolling window. The time of the minimum rolling-median HR was identified, and all raw HR measurements from the first available observation in the window through this minimum were retained. A linear regression model was then fit with HR as the dependent variable and elapsed time in hours as the independent variable. The resulting slope (in bpm per hour) was included in downstream analyses.

These daily wearable-derived features, consisting of daily step count, HR standard deviation, 95th percentile of HR, the daily HR and steps correlation, and the nocturnal HR slope from 8pm to the minimum HR, were aligned to each participant’s clinical (at times infection-related) alarm date. For each event, data were retained from 30 days before to 14 days after the alarm date. Repeated event-day values from the same participant and relative day were averaged within participant before participant-level analyses. To evaluate daily associations between activity and HR features while accounting for repeated measures within participants, linear mixed-effects models were fit at the participant-day level. Daily steps were the predictor and the HR feature was the outcome. Each was standardized using the mean and sample SD of the matched observations within the fitted analysis window. Models included a participant-specific random intercept and adjustment for standardized day relative to alarm and were fit by maximum likelihood. The primary model estimated the association between daily HR features and steps overall and within each analysis window. Model coefficients are reported as standardized beta estimates. P-values for the linear associations shown in **Figure 4a** were Holm-adjusted across the heatmap comparisons. For trajectory analyses, feature values were summarized across aligned days using centered 7-day rolling averages. In parallel, within-participant z-scored trajectories were computed to visualize deviations from each participant’s typical feature level. Window-level summaries were calculated by averaging each feature within each event and analysis window, followed by averaging repeated events within participant. Overall differences across the baseline, pre-alarm, and post-alarm windows were evaluated using Friedman tests, followed by paired Wilcoxon signed-rank tests with Holm adjustment for pairwise comparisons. Before event/participant averaging, outlier filtering was pooled within each feature over aligned event-day observations from days −30 to +14. Observations were retained when the absolute robust z-score, 0.6745 × (value − median)/MAD, was at most 6; the MAD filter was skipped when MAD was zero or nonfinite.

### Circadian and sleep-feature analyses

Cosinor models were applied to the HR data to extract the daily features of amplitude, acrophase, mesor, model R^2^, and p-value (testing non-zero amplitude) for each day and each participant, using the CosinorPy Python package. The SoundSleep Python package was used to extract sleep features for each participant-night, including wake after sleep onset as a percentage of total sleep time (WASO/TST) and number of awakenings. The daily circadian and sleep features were aligned to (potentially infection-related) clinical alarm events using the date on which the alarm was received by the study team. Plausibility constraints retained nonnegative amplitude, WASO/TST and awakening values, mesor values from 20 to 220 bpm, and R^2^ values from 0 to 1, inclusive. Within each feature, aligned event-day observations from days −30 through +14 were then retained when |0.6745 × (value − median)/MAD| was at most 6; the MAD filter was skipped for zero or nonfinite MAD. Acrophase was wrapped to [0,24) hours and excluded from MAD filtering. For window-based circadian analyses, the specified windows of baseline (days −30 to −15), pre-alarm (days −7 to −1), and post-alarm (days 0 to 7) were used. For each participant-event and feature, the mean value within the baseline window was subtracted from the mean value within each comparison window, yielding the feature change during the pre-alarm and post-alarm periods. Repeat events were averaged within participant before participant-level analyses. Changes were summarized separately by the antigen test outcome group. Overall differences across the three windows were assessed using Friedman tests, followed by paired Wilcoxon signed-rank tests with Holm adjustment for the three pairwise window comparisons. Acrophase used signed circular differences in [-12,12) hours and paired Wilcoxon tests with Holm adjustment; a Friedman test was not applied to clock time. The same applicable analyses were performed within antigen-test outcome groups and considered exploratory due to small subgroup sizes. For trajectory analyses, aligned daily cosinor features were summarized across all participants after averaging repeated events within participant by relative day. Mean values and standard errors were calculated for each day relative to day 0, and smoothed using a centered 7-day rolling window. Sleep was presented only as an illustrative case study. Participants with positive influenza or SARS-CoV-2 antigen outcomes were ranked first by the number of available sleep features and then by the number of observed feature values; this coverage rule selected a SARS-CoV-2 positive participant. Selection did not use the magnitude of the sleep change.

## Notes

### Competing Interest Statement

Michael P. Snyder reports cofounder and scientific advisory roles with Crosshair Therapeutics, Exposomics, Filtricine, Fodsel, iollo, InVu Health, January AI, Marble Therapeutics, Mirvie, Next Thought AI, Orange Street Ventures, Personalis, Protos Biologics, Qbio, RTHM, and SensOmics. His additional scientific advisory relationships include Abbratech, Applied Cognition, Enovone, Jupiter Therapeutics, M3 Helium, Mitrix, Neuvivo, Onza, Sigil Biosciences, TranscribeGlass, WndrHLTH, and Yuvan Research. He cofounded NiMo Therapeutics, has investment and scientific advisory relationships with R42 and Swaza, and holds an investment in Repair Biotechnologies.

### Author Declarations

The Institutional Review Board of Children's Hospital of Philadelphia gave ethical approval for this work (protocol 22-020440; parent study protocol 20-017872). The Institutional Review Board of Stanford University gave ethical approval for the Stanford component of this work (protocol 69457 and 60007).

