## Supplemental Information for "Real-time wearable detection of respiratory infections in pediatric transplant households"

SUPPLEMENTAL MATERIALS

**Supplemental Table 1. Participant-level wearable feature summaries across clinical alarm windows.** Baseline, pre-alarm, and post-alarm windows are days -30 to -15, -7 to -1, and 0 to +7, respectively. Red-alert rates are median [Q1, Q3]; other reported summaries are mean (SD).

| Feature | Baseline | Pre-alarm | Post-alarm | Overall P |
|---|---|---|---|---|
| Red-alert rate | 0.016 [0.000, 0.139] | 0.357 [0.286, 0.482] | 0.125 [0.000, 0.250] | 0.000843 |
| Median HR, bpm | 84.1 (9.5) | 86.4 (9.4) | 82.0 (11.0) | 0.683 |
| Minimum RHR, bpm | 59.3 (6.2) | 60.8 (7.5) | 60.1 (5.0) | 0.202 |
| 5th percentile HR, bpm | 65.7 (5.9) | 67.4 (5.6) | 66.4 (6.3) | 0.269 |
| Estimated HR at zero steps, bpm | 77.7 (7.9) | 79.5 (6.2) | 76.9 (6.6) | 0.0111 |
| Within-day HR SD, bpm | 15.9 (3.5) | 15.4 (4.2) | 14.3 (4.2) | 0.319 |
| 95th percentile HR, bpm | 113.8 (12.5) | 114.4 (14.1) | 108.9 (15.7) | 0.829 |
| Within-day HR-steps correlation | 0.525 (0.142) | 0.504 (0.150) | 0.520 (0.108) | 0.931 |
| Nocturnal HR slope, bpm/hour | -2.715 (1.490) | -3.102 (1.519) | -1.999 (2.308) | 0.223 |
| Cosinor amplitude, bpm | 16.9 (5.6) | 16.4 (7.8) | 16.5 (8.0) | 0.646 |
| Cosinor mesor, bpm | 83.2 (8.3) | 84.7 (8.4) | 82.3 (12.0) | 0.0284 |
| Cosinor model fit, $R^2$ | 0.290 (0.120) | 0.269 (0.126) | 0.253 (0.119) | 0.099 |

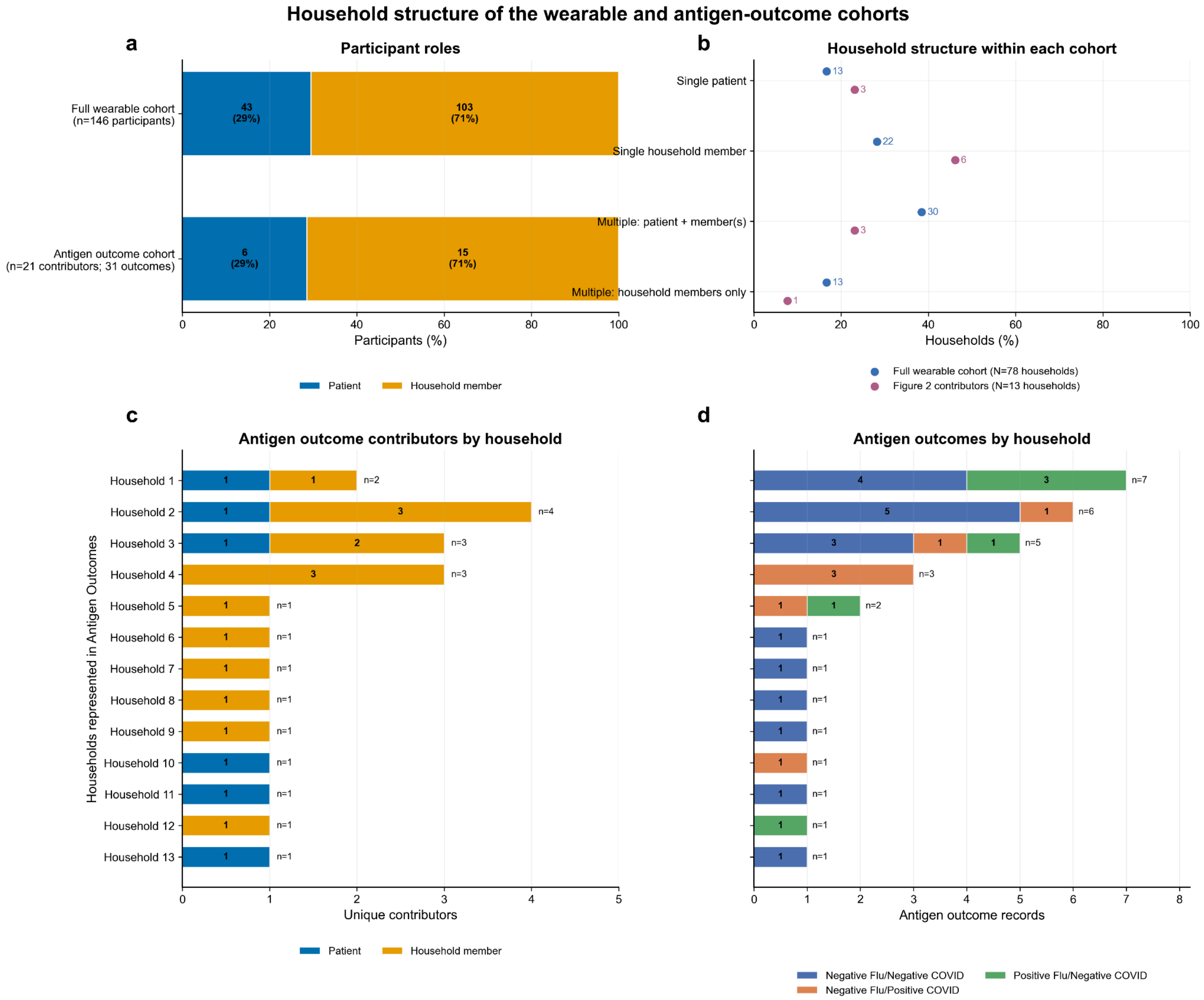


**Supplemental Figure 1: Household structure of the wearable and antigen-outcome cohorts. a**, Proportions of patients and household members among the full wearable cohort and the 21 participants contributing to the 31 antigen test outcomes. **b**, Household composition within the full wearable cohort and the antigen outcome cohort. Households were categorized as containing a single patient, a single household member, multiple participants including a patient and one or more household members, or multiple household members without a patient. Numbers beside each point indicate the number of households in the corresponding category. **c**, Numbers and roles of unique antigen outcome cohort contributors within each of the 13 represented households. **d**, Numbers and antigen test outcomes of records contributed by each household.

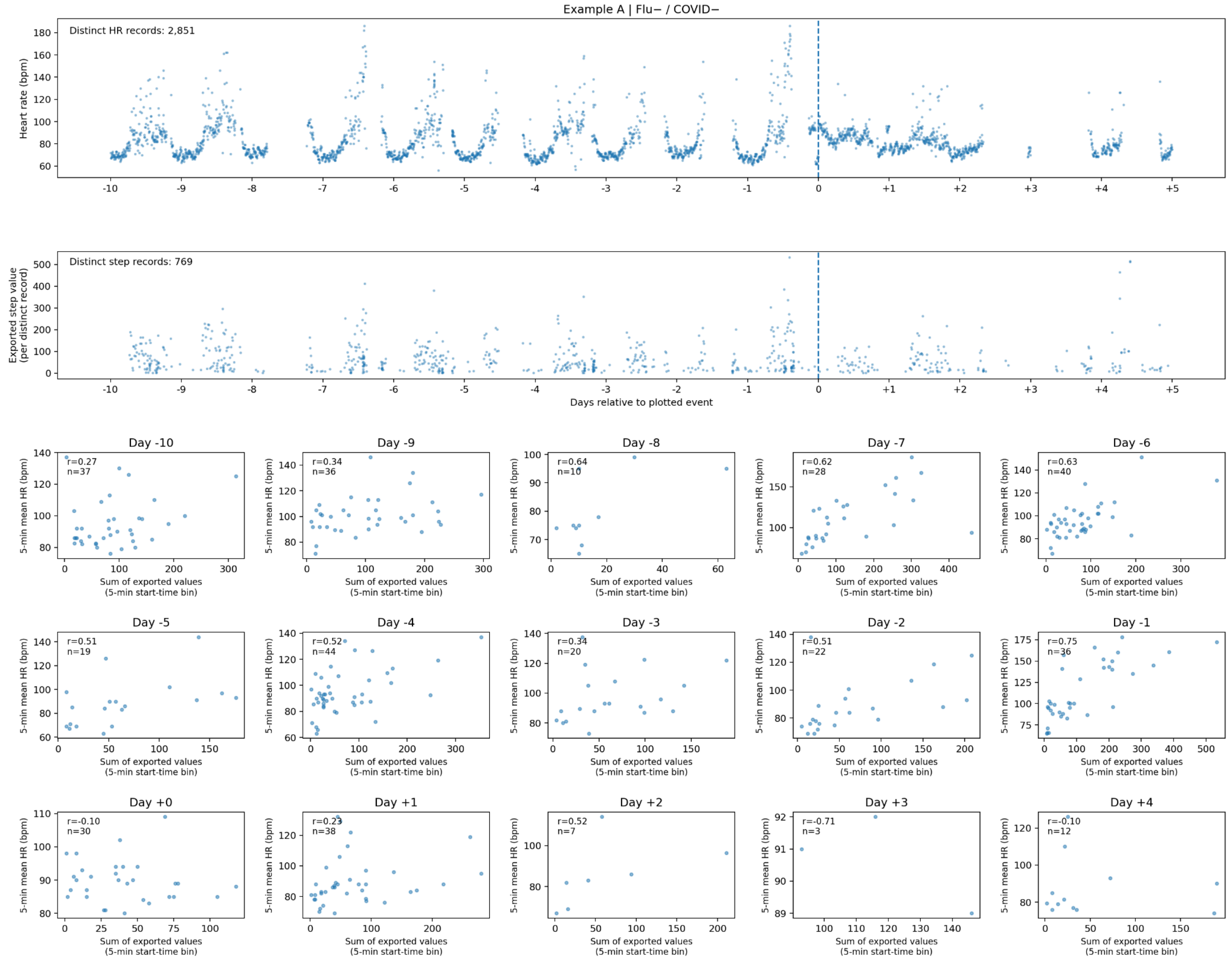


**Supplemental Figure 2: Descriptive wearable data example for a Negative Flu/Negative COVID participant.** Dates are shown relative to the plotted event (day 0; dashed line). The top two rows show HR and steps values after exact repeated records were removed. Each lower-panel point combines mean HR with the sum of step values assigned by the start time of the record to the same five-minute bin; only bins containing both record types are shown. n denotes observed bin pairs, and r is the descriptive Pearson correlation, shown for at least three pairs with variation in both variables.

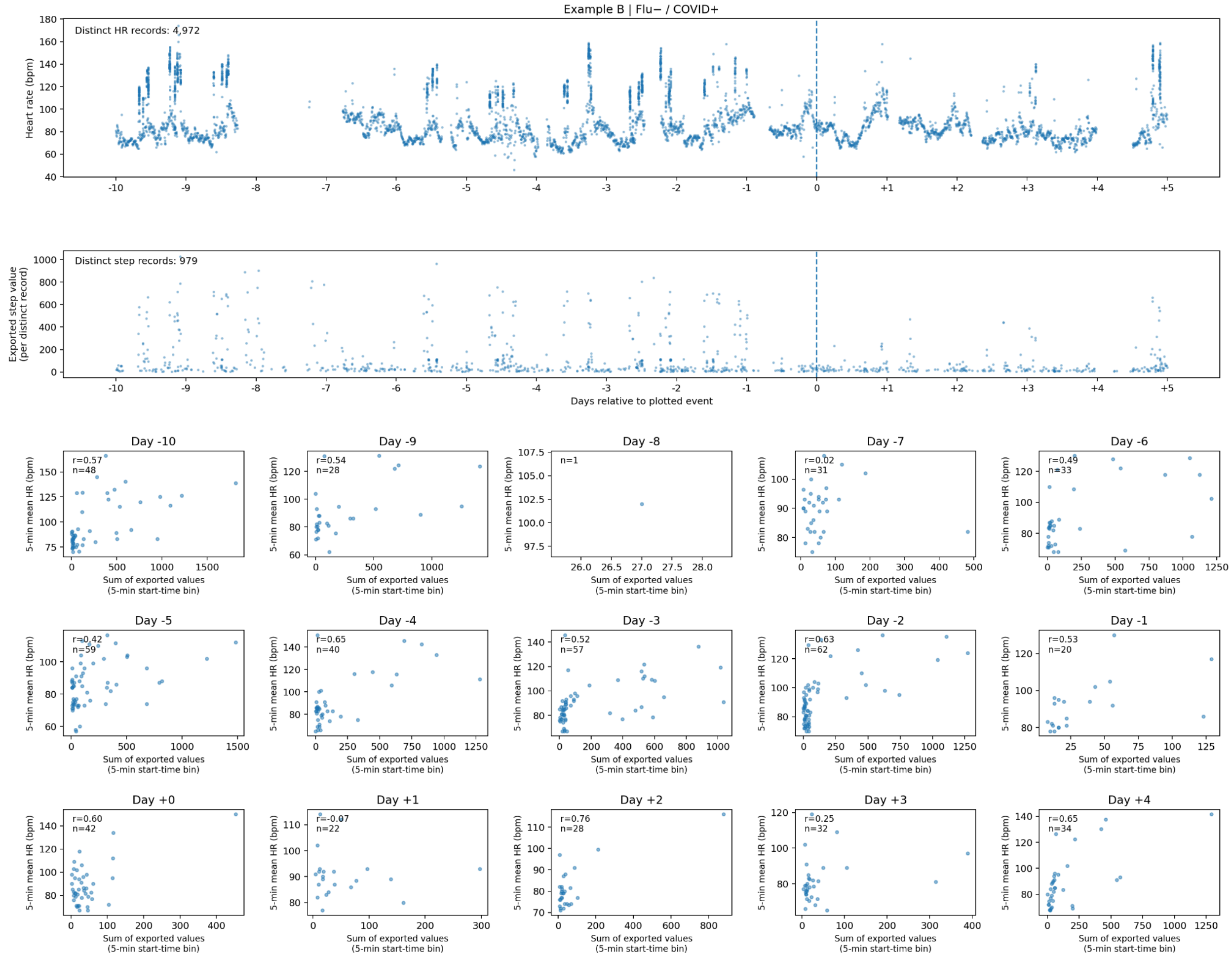


**Supplemental Figure 3: Descriptive wearable data example for a Negative Flu/Positive COVID participant.** Dates are shown relative to the plotted event (day 0; dashed line). The top two rows show HR and steps values after exact repeated records were removed. Each lower-panel point combines mean HR with the sum of step values assigned by the start time of the record to the same five-minute bin; only bins containing both record types are shown. n denotes observed bin pairs, and r is the descriptive Pearson correlation, shown for at least three pairs with variation in both variables.
